# Human agency and infection rates: implications for social distancing during epidemics

**DOI:** 10.1101/2020.04.11.20062042

**Authors:** Christopher Bronk Ramsey

## Abstract

Social distancing is an important measure in controlling epidemics. This paper presents a simple theoretical model focussed on the implications of the wide range in interaction rates between individuals, both within the workplace and in the social setting. The model is based on well-mixed populations and so is not intended for studying geographic spread. The model shows that epidemic growth rate is largely determined by the upper interactivity quantiles of society, implying that the most efficient methods of epidemic control are interaction capping approaches rather than overall reductions in interaction.

The theoretical model can also be applied to look at the dynamics of epidemic progression under various scenarios. The theoretical model suggests that with no intervention herd immunity would be achieved with a lower overall infection rate than if variation in interaction rate is ignored, because by this stage almost all the most interactive members of society would have had the infection; however the overall mortality with such an approach is very high. Scenarios for mitigation and suppression suggest that, by using interactivity capping, it should be possible to control an epidemic without extreme sanctions on the majority of the population if *R*0 of the uncontrolled infection is 2.4. However to control the infection rate to a specific level will always require the switching on and off of measures and for this reason eradication is likely to be a less costly policy in the long run. While social distancing alone can be used for eradication it is not be a good mechanism on its own to prevent reinfection. The use of robust testing, quarantining, and contact tracing would strengthen any social distancing measures, speed up eradication, and be a better tool for prevention of infection or reinfection.

## 1 Introduction

The use of social distancing has become a critical one for countries attempting to deal with the outbreak of COVID-19[1]. The difficulty is particularly in coming up with strategies which are sustainable in the long-run, or which are capable of eliminating the virus entirely. This paper sets out to explore one aspect of this question with a very simple epidemic progression model focussed on looking at the implications of variability in individual contributions to the spread capacity.

The key parameter for understanding the epidemic dynamics is *R*0, which is the average number of other individuals infected by someone who carries the disease when people are operating normally. If this figure is above 1, then exponential growth is expected until there is saturation due to some form of herd immunity. If the figure is less than 1 then the disease will, in time, be eliminated. As *R*0 is the initial rate, at this point it is assumed that overall infection rates are low and both immunity and the probability of passing on the infection to someone already infected can be ignored.

In many ways, the best approach is to use large-scale epidemic simulation [2, 1] or to use a complex network model [3], but it is worth testing these approaches against simpler models which can be used to explore aspects of the problem. In this paper, a totally theoretical probability-based approach is taken, which treats the population as a whole, and which specifically addresses variation in social interaction of individuals, with the aim of understanding how different human agents contribute to the spread. This model is not useful for studying geographic spread and, because it does not have any regional granularity, it is cannot address specific questions of how to deal with households or institutions, though in principle the model could be considered for institutions as approximately independent units, where infection rates are low. The purpose of the model is not to replace fuller simulation or network based studies but to provide a model which is simple enough for non-specialists to use and which can be used to understand why some strategies might be more effective than others.

## 2 A simplified theoretical model

Part of the motivation for this theoretical approach are the known limitations in the types of social contact data which underly simulation models, and are required for quantifying networks. In particular, the types of survey data used may under-report chance interactions [4], and indeed the scale of variation in the length, intensity and closeness of contacts is hard to elicit from such data. The amount of interaction any individual has in a complex modern society will vary over a very wide scale, and probably over a wider scale than contact survey data would suggest. This is partly due to the upper limit on the number of contacts reported, either because the respondent cannot remember them, or because they do not necessarily think they are significant. For the purposes of this model the number of contacts is, in any case, not as significant as total one-one contact time for virus transmission.

Here likely ranges of one-one contact times are estimated, and simplified model presented which only uses this parameter to model infection spread within the population.

### 2.1 A stepwise approach to epidemic progression

The parameter *R*0, if considered as a global model parameter, is the average expected number of new infections arising from each randomly selected infected individual from an overall population of N. In a stepwise process, the probability that each individual *i* is infected in cycle *j* can be defined as *p*_*ij*_ with and the probability that they will transmit the infection on as *t*_*i*_; this is a measure of how intensely they interact with others in society as a result of their life choices and occupation. From this it is possible can extract the expected number of infections at one iteration of the cycle *M*_*j*_ and the number in the next cycle *M*_*j*+1_.

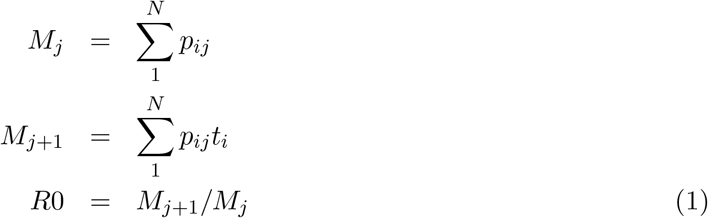

### 2.2 Human interaction sensitivity

In considering the effect of individuals within the population, it is important to note that *p*_*i*_*j* and *t*_*i*_ are likely to be very highly correlated. This is because a person who has a high probability of passing an infection on is also much more likely to pick it up. A realistic assumption for air-borne viruses would be that these two are actually directly proportional because they both arise from the amount of time spent in close proximity to others. This can be used to derive a value for *R*0 from the average transmissibility as 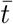:

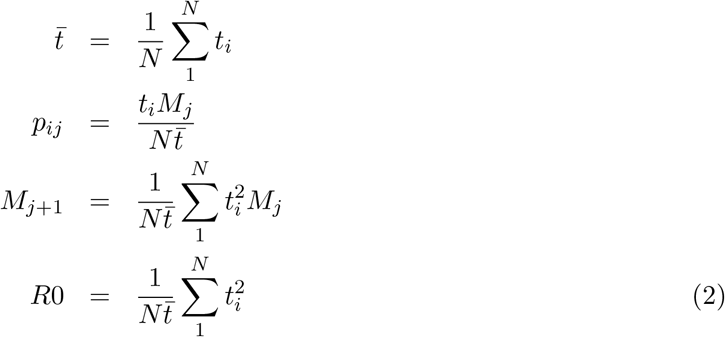

This simple model, with its non-linear dependency on 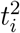, suggests that the *R*0 parameter for the population will be dominated by the contribution from those with high *t* values.

### 2.3 Variability in interactivity

The parameter *t* for each individual is likely to be very variable within a population and indeed the exact numbers will vary between countries and regions. There is some good data on contact numbers and intensity[4], but these are not in exactly the form needed for this model. However, some reasonable upper and lower limits can be estimated. At the upper limit will be those who spend most of their day in close proximity to four other individuals in crowded spaces (*ca*. 50 person hrs/day), and at the other end of the scale, people who only have close interaction very rarely when doing essential errands, or those living alone who are rarely visited (*ca*. 0.5 person hrs/day). Whether contact within the household is considered is a difficult question, because households are particularly integrated. It might be better to consider each household as a super-individual - so with extra interactions and greater chances of picking up the infection together; this issue will not be addressed here, but obviously is within the simulation studies. The exact numbers are not important but the variability is, and here it is assumed that there is likely to be a 95% probability range of about two orders of magnitude in *t*. The distribution *f* (*t*) will be estimated to be log normal (using the estimates above, the median would be *ca*. 5 person hrs/day).

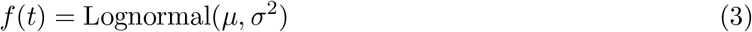

Integrating over the whole population gives:

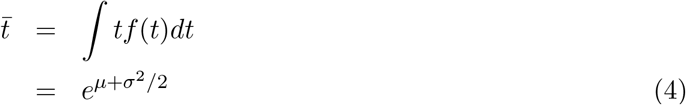

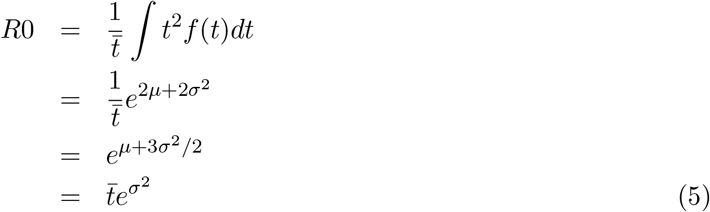

This leads to the expected result that, if all individuals in the population have the same *t* value (*σ* = 0), this will also be *R*0 for the population. However, if the 95% probability range for *t* covers a range *α* then *σ* = ln *α/*4, implying that with *α* = 100 as estimated above, 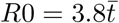. While this is numerically correct, the very strong weighting to high *t* values makes this highly dependent on the upper limit of interactivity, and because there are practical limits to this, if the upper value of t is capped at the 2*σ* level this is reduced to 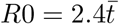which may be more realistic. For comparison if *α* is only 10 then 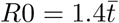, whether or not a cap for a reasonable upper limit is used.

## 3 Implications of the theoretical model framework

The analysis above shows the dominant effect of the upper end of the *t* distribution on *R*0 and implies that an effective way to reduce the growth rate to some lower effective value (*R*_eff_) is to reduce *σ* rather than reducing 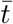, or more specifically to reduce the interactivity of those members of the population with the highest values of *t*.

### 3.1 Overall reductions in interaction

One way to reduce *R*_eff_ is to reduce the interaction level of the entire population by the same factor. It can be seen from equation 5 that this simply requires a reduction in 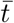 of the *R*_eff_ */R*0. For example to reduce an *R*0 of 2.4, which is probably a reasonable estimate for COVID-19[1], to *R*_eff_ *<* 1, requires a drop in interactions of a factor of more than 2.4. This is probably very difficult for those with lower interaction levels to achieve and is almost certainly not the most efficient way to do this.

### 3.2 Partitioning of interaction

Perhaps the most interesting impact of the high correlation between *p*_*ij*_ and *t*_*i*_ is the effect on the partitioning of interaction between individuals. This can also be seen directly from equation 5, because a more even partitioning of interactions between individuals will have a significant effect on *R*_eff_. In the limit that all interactions are evenly distributed through the population, it might be possible to reduce *R*_eff_ by the factor of 2.4 while keeping the overall interaction levels (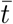) the same. This is not going to be practical but it underlines the important of considering partitioning in designing distancing strategies.

The same point can be illustrated by a simple thought experiment. There is an activity which involves a high interaction level and two people who could be given that task, who otherwise would be relatively isolated. If one person works and the other is completely isolated, the one working has both a high probability of infection and a high probability of passing that on. If the job is divided between the two people, their infection rates are both half that of the individual worker but their probability of passing the infection on is also halved, and so the overall effect on propagating the disease is halved.

### 3.3 Proportions of the population contribution to *R*0

Another way to look at this is to consider the effect of different sections of the *f* (*t*) distribution on *R*0. If the log normal distribution truncated at +2*σ* is used, as discussed above, 50% of the value of *R*0 is generated by the most interactive 5% of the population. Likewise the bottom 50% of the population in interaction terms only contributes about 2% to the total, which makes it clear why modifying their behaviour is unlikely to have significant effect: these are people who are unlikely to get the infection and unlikely to pass it on.

### 3.4 Targeting of interventions

The most efficient way to implement social distancing measures is to target them at the most active members of the population. Indeed, as seen above in the discussion of partitioning, the key thing is to limit the maximum amount of interaction any individual has. Individuals with very high interaction levels during an epidemic present a risk both to themselves and to the population as a whole. This is well known from anecdotal discussion of super-spreaders, but is numerically clear from equation 2.

This is exactly what you would expect from network analysis [3], where immunisation of high degree nodes is shown to be far more effective than random immunisation. Essentially this is the same strategy, except here it is proposed that reduced interaction of highly interactive individuals is used where immunisation is not available.

Conversely the section of the population which is least interactive has almost no influence on *R*_eff_ at all. It is clear from this analysis, if not already, that trying to drive down interaction levels that are already far below the median will have almost no effect on *R*_eff_. However, any reduction in interaction will reduce the risk of infection for that individual and so may still be a useful measure, especially for those at greatest risk from the infection.

Putting some numbers on this from the estimates above, if the maximum interaction any individual can have is limited to 1*σ* above the median level in the log normal distribution this will reduce *R*_eff_ by a factor of 0.51, so almost enough on its own to reduce the an *R*0 of 2.4 to an *R*_eff_ of 1. If the median contact was 5 person hrs/day this would imply an upper limit of 15.8 person hrs/day. A limit set at the median value would reduce *R*_eff_ by a factor of 0.20, certainly low enough to eliminate the disease, and somewhere between these two would probably be the optimum (see scenarios below).

These estimates are based on a whole population group, and more granular information could be used to inform *p*_*ij*_, since this will also depend on local infection rates over time: it would make sense to use local case rates rather than just national. Another key thing to consider is that it is the average interaction rate over a time period relevant to the incubation that is relevant. Temporary high interaction levels, for example at a single event, will be strongly mitigated if combined with the periods before and after have much lower interaction rates. On the other hand, residential events with intense interaction over several days will probably have the greatest impact on raising *R*_eff_.

### 3.5 Contact tracing, testing and immunisation

This analysis also has implications for contact tracing[5], because it shows that the people for whom this is most important are those who interact most strongly. Thus if it can be made to work efficiently on this group alone, it could help to limit the effect of the highly active groups on the overall statistics. Likewise, if it is not possible to reduce interaction rates (for front-line medical staff for example), then the risks of passing infections on can be reduced by other measures such as personal protection and regular testing. It is likely that contact tracing and regular testing of the least interactive portion of the population would be less likely to have a major impact.

In a period when there are still significant risks, one way to allow interaction risk events to proceed, and institutions to remain open, would be to make use of contact tracing and testing to ensure that *p*_*ij*_ is low for all individuals at the start.

There are also other synergies here with contact tracing strategies, and in particular with the use of mobile phone apps for implementation of this. These apps would be ideal for alerting individuals that their overall interaction was high so that they can socially distance for a while to reduce this. They would also be an ideal way of identifying those individuals who would most benefit from immunisation when and if that becomes available[3].

## 4 Scenario modelling

### 4.1 Dynamic modelling

The above framework has been developed into a simple dynamic step-wise model. The population is divided into 800 quantiles, each with a different *t* value, covering *±*4*σ*. Initially these are seeded with a probability of infection set for the particular model run. Using the *t* values for each quantile the expected number of potential infections can be calculated directly for the population as a whole. These are then partitioned to the population in proportion to the *t* values of all individuals. The actual infection probability for each quantile is then reduced by the proportion of that quantile which has already been infected.

Unless otherwise stated an *R*0 of 2.4 will be used, with a 95% range of interactivity of 100 and a maximum interaction rate set at 10 time the median. This gives a 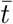 of 0.995 and a median value of *t* of 0.558. All outputs are expressed in cycles of infection, following the logic of this simple model; in the case of COVID-19 this is expected to correspond roughly to weeks. In practice with a distributed population, everything would be expected to lag more than this model would suggest. It is also important to note that the parameter modelled here is new infections, symptoms will lag this and detected cases (especially if they are on hospitalisation) will lag further, and deaths, further still. This model is best suited to looking at endemic infection within a single integrated population. The initial seed infection rate probability is chosen to be 0.0001 which is the level at which it is very obvious you have an issue; the seed infection is assumed to be distributed in proportion to individuals interactivity which fits with the overall logic of this model.

For many of the scenarios it is useful to further split the population into groups with different interventions. For those cases, in this paper three groups have been used: 60% in group 1, 20% in group 2, which is assumed to be vulnerable, and 20% in group 3, which are assumed to be key workers and therefore not subject to some sanctions. Where mortality is discussed, this is assumed to be 0.5% for groups 1 and 3, and 5% for group 2; this gives an IFR of 1.4% overall which is the upper limit of what is expected[6].

For scenario testing, threshold levels and a delay response can be set in the model. The levels which have been chosen for the scenarios below (where relevant) are 0.0002 (double the seed probability), 0.0004 and 0.001. The response time is 3 cycles which is probably the minimum practical, given that cases have to be detected and some notice of measures has to be given. Longer response times have fairly predicable effects, particularly in increasing the case load in the initial pulse.

In general four different levels of response using two different strategies have been considered. The first strategy, based on the analysis above is to limit the interactivity of the most interactive individuals, essentially capping this at a particular point. The second strategy is to reduce everyone’s activity by the same factor. In practice most policy interventions are likely to be a mixture of these. The four levels of interaction considered are:

1. Limited Distancing: this entails a capping of activity at the +1*σ* level, which with the param-eters 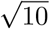 above would entail 10 above the median level of interaction (*ca* 15 person hrs/day).An approximately similar effect can come from reducing the whole population’s activity by a factor of 0.5.
2. Enhanced distancing: this entails a capping of activity at the +0.5*σ* level, which with the parameters above would entail 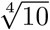 above the median level of interaction (*ca* 9 person hrs/day). An approximately similar effect can come from reducing the whole population’s activity by a factor of 0.33.
3. Strong distancing: this entails a capping of activity at the median level, which with the parameters above would entail at the median level of interaction (*ca* 5 person hrs/day). An approximately similar effect can come from reducing the whole population’s activity by a factor of 0.2.
4. Isolation: this entails a capping of activity at the *−*1*σ* level, which with the parameters above would entail 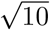 below the median level of interaction (*ca* 1-2 person hrs/day). It is hard to replicate this with population-wide reductions in activity, here a factor of 0.1 is used (the estimated ratio required is about 0.07).

These levels are chosen specifically because limited distancing results in an *R*_eff_ just above 1 and enhanced distancing results in an *R*_eff_ of just below 1, which allows for control of the epidemic progression. It is helpful to consider caps in terms of natural variation in activity since it should be possible for people to reduce their interaction levels to the median level, since half the population does this anyway. It is much harder to assess whether everyone reducing their interaction level by a factor of over 2 is really practical.

Many of the findings of the scenarios here will be similar under other models, the key here is control of *R*_eff_. However, the actual dynamics and mortality rates are model dependent and, as this is a purely theoretical model, should not be treated as predictions of what would happen in practice. Further simulation models should be used to test the implications presented here.

### 4.2 No intervention

One possible scenario is that there is no intervention. In this case the model runs until herd immunity is reached (see Figure 1). The peak in new cases occurs around cycle 9, with about half the overall cases within three or four cycles. In this model herd immunity is reached with an overall infection rate of 41%. This is lower than the 81% estimated in the simulation model[1], which could be due to limitations of the model here, or it could be due to underestimation of interactive variability in the simulation. The herd immunity level is dependent on the variability estimates, but even with a factor of only 10 it is still 73%, and to get to an estimate of 81%, it is necessary to lower this to a factor of only 3, which is unrealistic.

**Figure 1:**
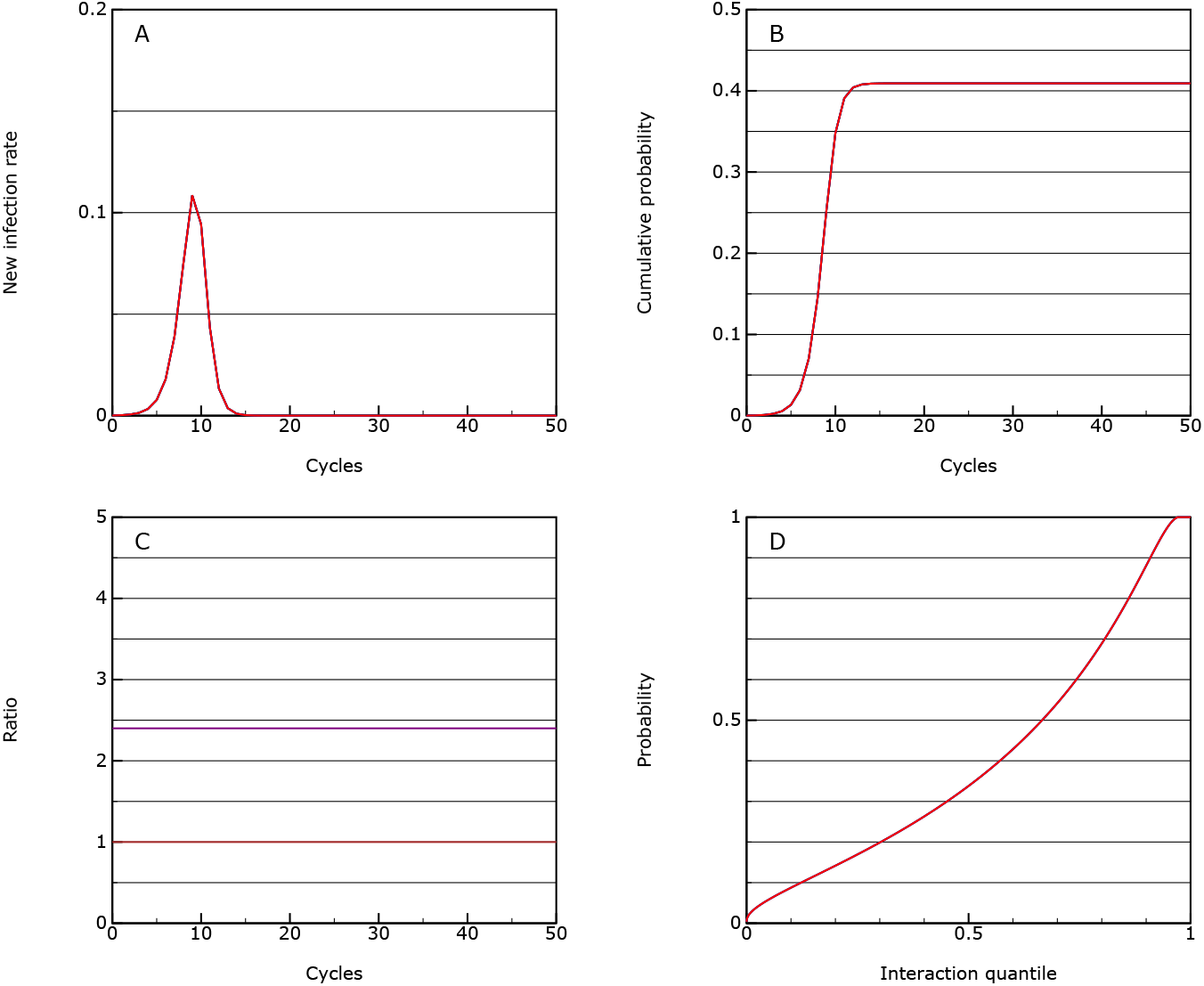
This shows the model behaviour with no intervention; A: the rate of new infections as a proportion of the population in each cycle of the epidemic; B: the cumulative infection rate for the population; C: the *R*_eff_ value (purple) which in this case is equal to *R*0, and the allowed interaction rate compared to normal (brown), through the epidemic; D: the accumulated infection probabilities for quantiles of the population ordered by interactivity.

The overall mortality rate is 0.0057, assuming that overload in the health systems does not raise the prior assumptions about mortality rate.

The levels required for herd immunity here are due to the treatment of quantiles with different activity levels. In this scenario the very active individuals are almost all infected (Figure 1D) and thereby reduce the overall rate of rise for the population as a whole, until *R*_eff_ falls below 1. This is something which needs further investigation within the simulation studies, and the data from the epidemic.

### 4.3 Protection

This is the same as the no intervention scenario but introduces strong distancing for the vulnerable group at a case threshold of 0.0002 and isolation at 0.0004. Under this model the overall progression of the epidemic is similar but the mortality rate drops by a factor of about 2 to 0.0027 (see Figure 2).

**Figure 2:**
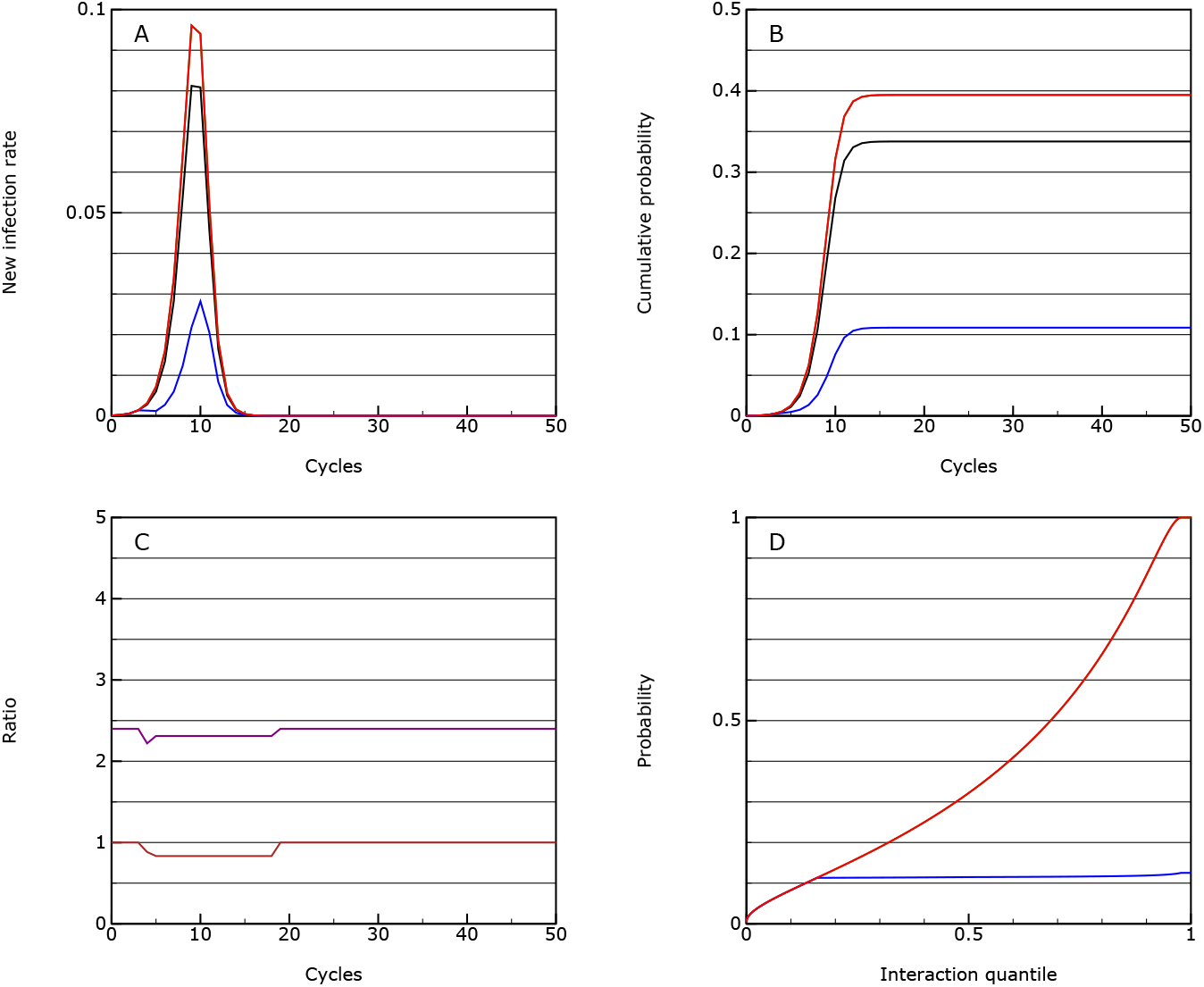
As for Figure 1, but where the vulnerable group (shown in blue), is protected through partial isolated from the rest of the population; the population average is shown in black.

### 4.4 Mitigation

One possible option considered is to slow the peak, but this turns out to be quite difficult to achieve in this model. It requires you to choose those interventions which reduce *R*_eff_ to just around 1 which is very hard to do. Here it is achieved by setting the following limits at the three thresholds:

- 0.0002: the vulnerable group goes onto strong distancing while all others go onto limited distancing.
- 0.0004: the vulnerable group goes into isolation
- 0.001: the main group goes into enhanced distancing

This is just enough to control the disease at a new case rate of about 0.001 (see Figure 3). However the controls have to be cycled on and off for the main group.

**Figure 3:**
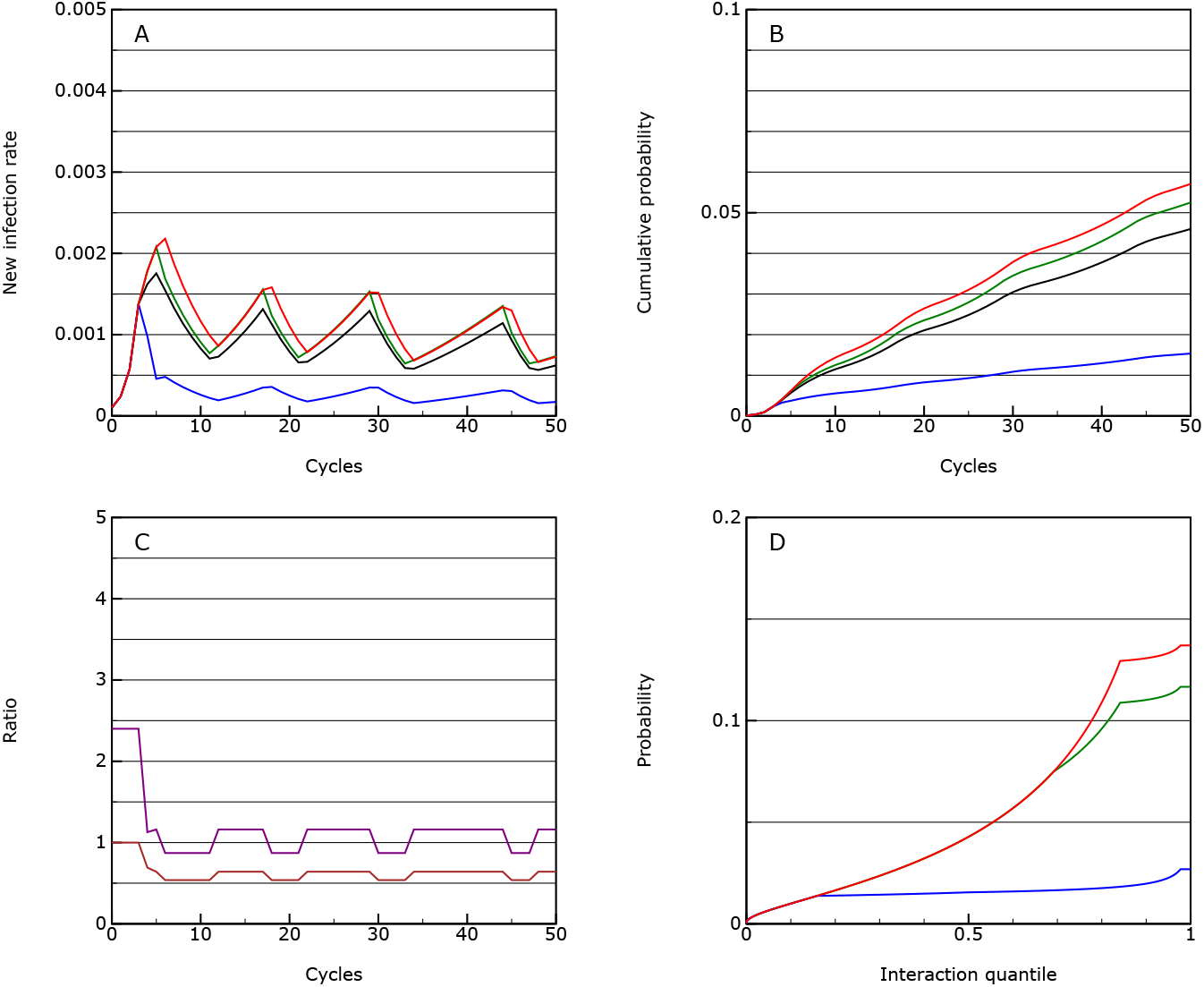
As for Figure 1, but where the vulnerable group (shown in blue), is partially isolated from the rest of the population, the majority of the population (green) has significant distancing measures applied and key workers (red) have limited distancing measures; the population average is shown in black; this is the mitigation scenario.

The mortality over 50 cycles is 0.00037, so almost an order or magnitude lower than the protection scenario. However, the infection will keep on flaring up and if taken over 10 years, the overall mortality may even rise above the protection scenario.

The overall level of interaction between people in the population as a whole is reduced by a factor of 0.63, if activity capping is used and by 0.43 if an activity factor is used. This pattern is replicated with many of the other scenarios, with the societal cost being far greater if overall activity factors are used rather than caps to activity levels.

### 4.5 Suppression

Given that it is possible to bring *R*_eff_ to below 1, the next scenario looks at sustaining this for longer to keep the overall case rate lower. This is achieved using sanctions at the following limits:

- 0.0002: the vulnerable group goes onto strong distancing while all others go onto limited distancing.
- 0.0004: the vulnerable group goes into isolation and the main group goes into enhanced distancing.
- 0.001: the main group goes into strong distancing

As expected this essentially behaves similarly to the mitigation strategy but controls around the lower threshold value of 0.0004 (see Figure 4), and indeed, given the nature of the response in the model, it should be possible to stabilise at any level using this methodology (two levels of response to give a value of *R*_eff_ just below and just above 1).

**Figure 4:**
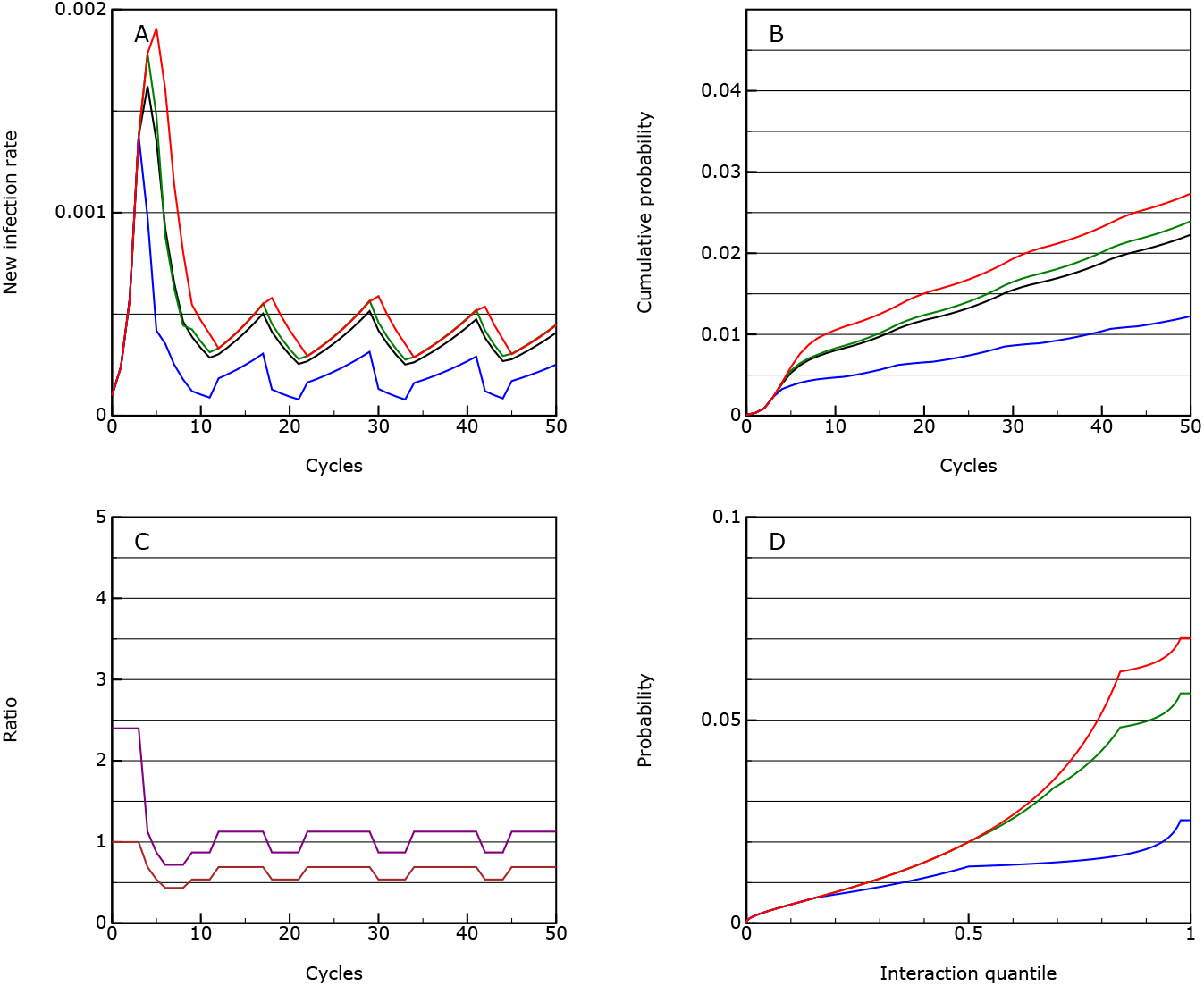
As for Figure 1, but with lower thresholds for measures to be applied; this is the suppression scenario.

The mortality over 50 cycles is 0.00022 with much of this being in the first pulse.

The overall level of interaction between people in the population as a whole is reduced by a factor of 0.65, if activity capping is used and by 0.43 if an activity factor is used, essentially the same as in the mitigation scenario. There is no real penalty in stabilising the infection rate at a lower threshold.

### 4.6 Lockdown

So far the scenarios have not involved putting the main group under very severe restrictions, because, according to this theoretical model, this should not be necessary to bring *R*_eff_ below 1 if capping of activity levels is used. However, it is worth seeing the effect of more severe restrictions is under this model and here is achieved using sanctions at the following limits:

- 0.0002: the vulnerable group goes onto strong distancing while all others go onto limited distancing.
- 0.0004: the vulnerable group goes into isolation and the main group goes into strong distancing.
- 0.001: the main group goes into isolation

This leads to a less stable case level, and ends up with cycling between all three states (see Figure 5). This is because *R*_eff_ is less than one in the all but the lowest state. The overall activity levels remain around 0.65 (or 0.46 if a factor approach is used), but the activity levels fluctuate in a way which is likely to be very disruptive. The mortality over 50 cycles is similar at 0.00032 (if anything higher because sometimes all restrictions are lifted) and there seems to be little to be gained from this strategy from a technical point of view, but may be necessary politically if other attempts at reducing interaction levels do not work.

**Figure 5:**
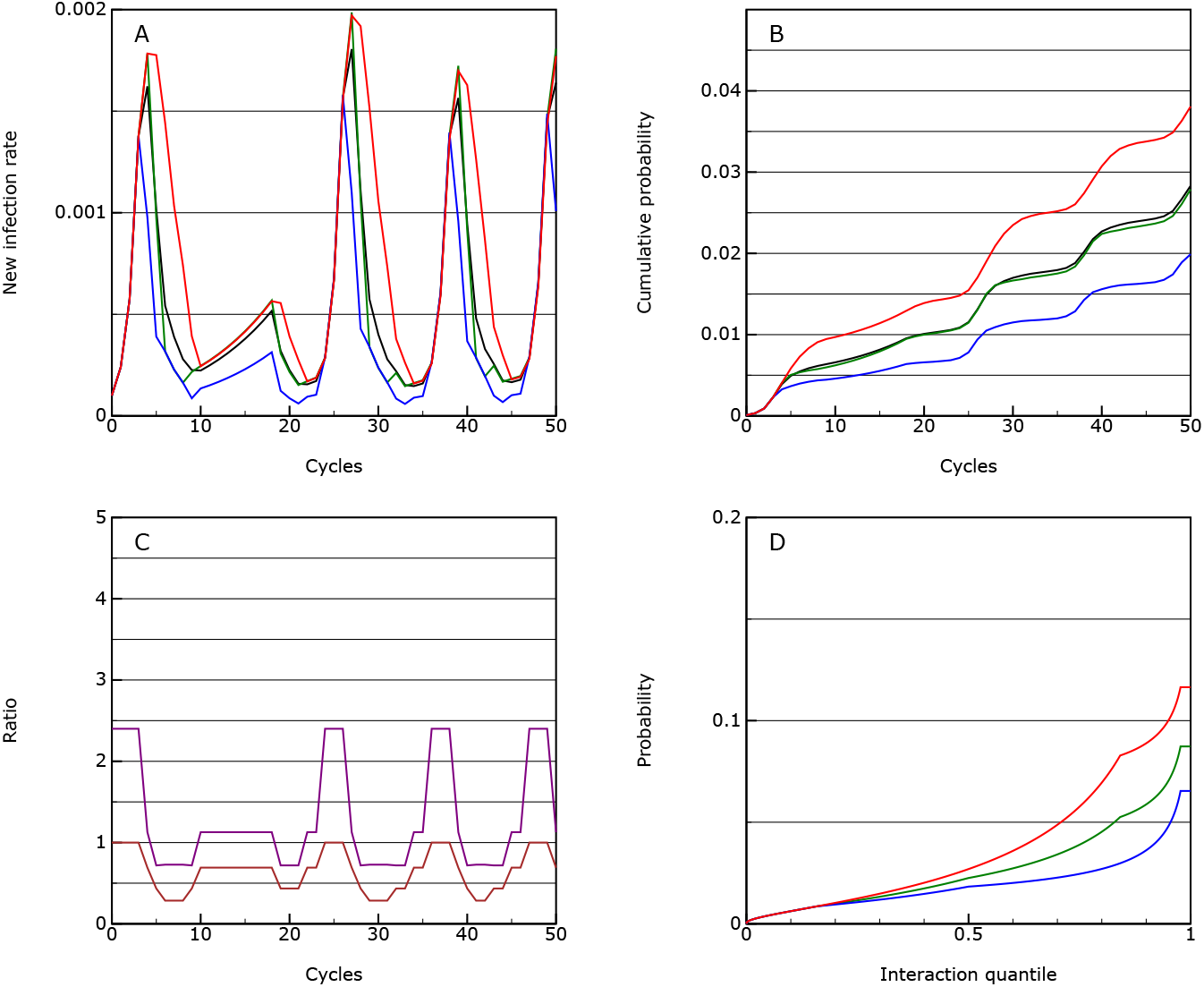
As for Figure 4, but with more extreme measures imposed on the majority of the population; this is the lockdown scenario.

Interestingly if no reduction in activity of the key worker group is made, it is very difficult to stabilise the situation, even with the majority of the population in lockdown (see Figure 6). If they remain at normal interaction levels, the epidemic will take off in that group and spread to the rest, with over 20% of this group infected by the end (and almost all of the most interactive individuals). The overall mortality rate is much higher at about 0.001 with much of that, inevitably, in the key worker group. This finding is also true under the mitigation and suppression strategies and underlines the need for some social distancing for this group (combined with personal protection and testing). The main risk in managing an epidemic, especially if initial measures are not brought in fast enough, is that the key workers become over-stretched and if anything their interaction levels rise: this positive feedback effect is something to be avoided if at all possible.

**Figure 6:**
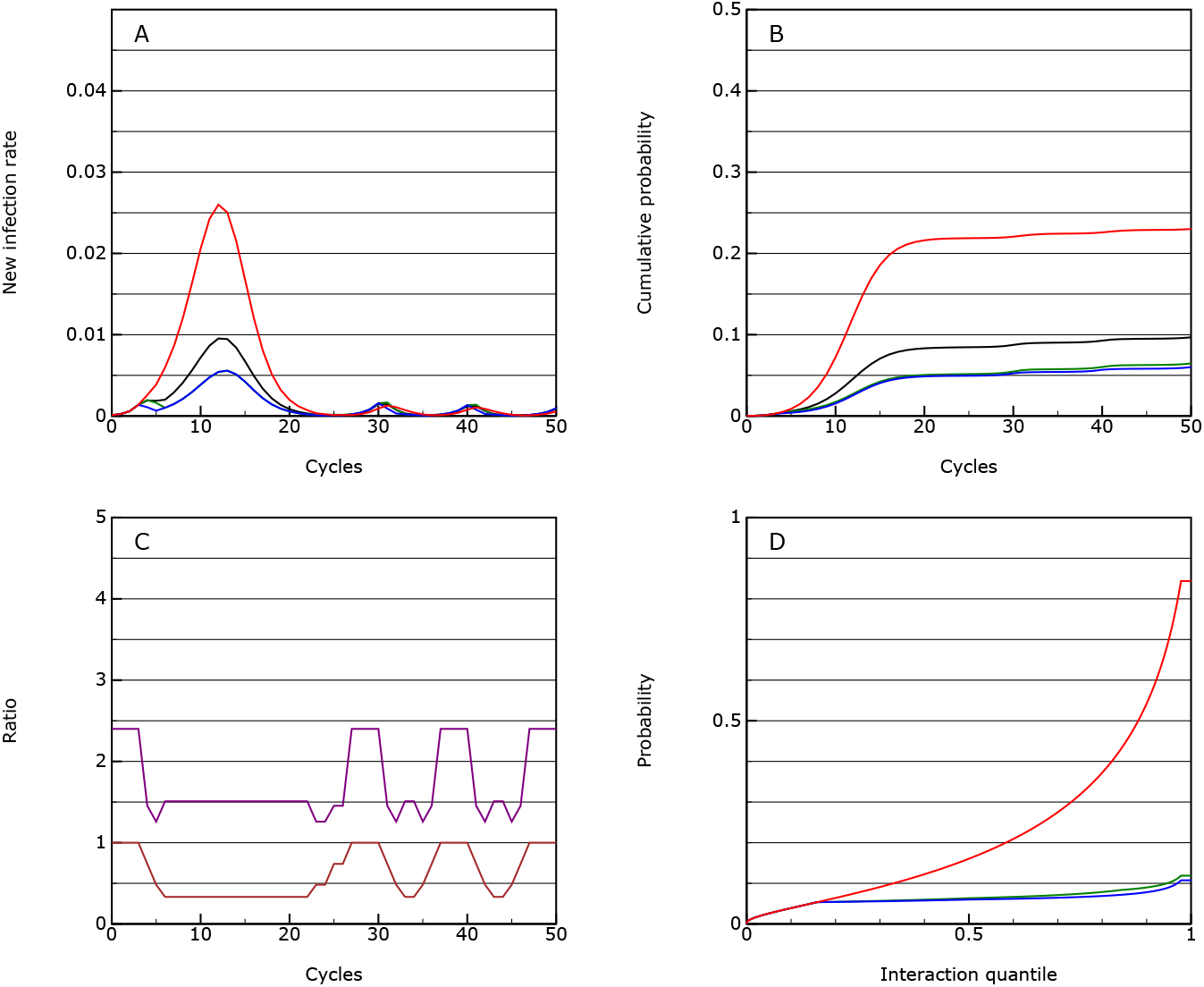
As for Figure 5, but with the key worker group maintaining their normal interaction levels.

### 4.7 Eradication

Give it is possible to reduce *R*_eff_ below 1, it is clearly possible to go for eradication. This can be achieved by lowering the first threshold to zero (permanent sanctions). The measures are then set at the following levels:

- 0.0000: the vulnerable group goes onto strong distancing, the main group goes into enhanced distancing and the key workers go onto limited distancing.
- 0.0004: the vulnerable group goes into isolation.
- 0.001: the main group goes into strong distancing.

This will effectively eradicate the disease (case rate to less than 1 in a million) over 50 cycles (see Figure 7). Clearly stronger sanctions would do so faster, but actually if these are not applied to the key worker group too, the differences are not that great.

**Figure 7:**
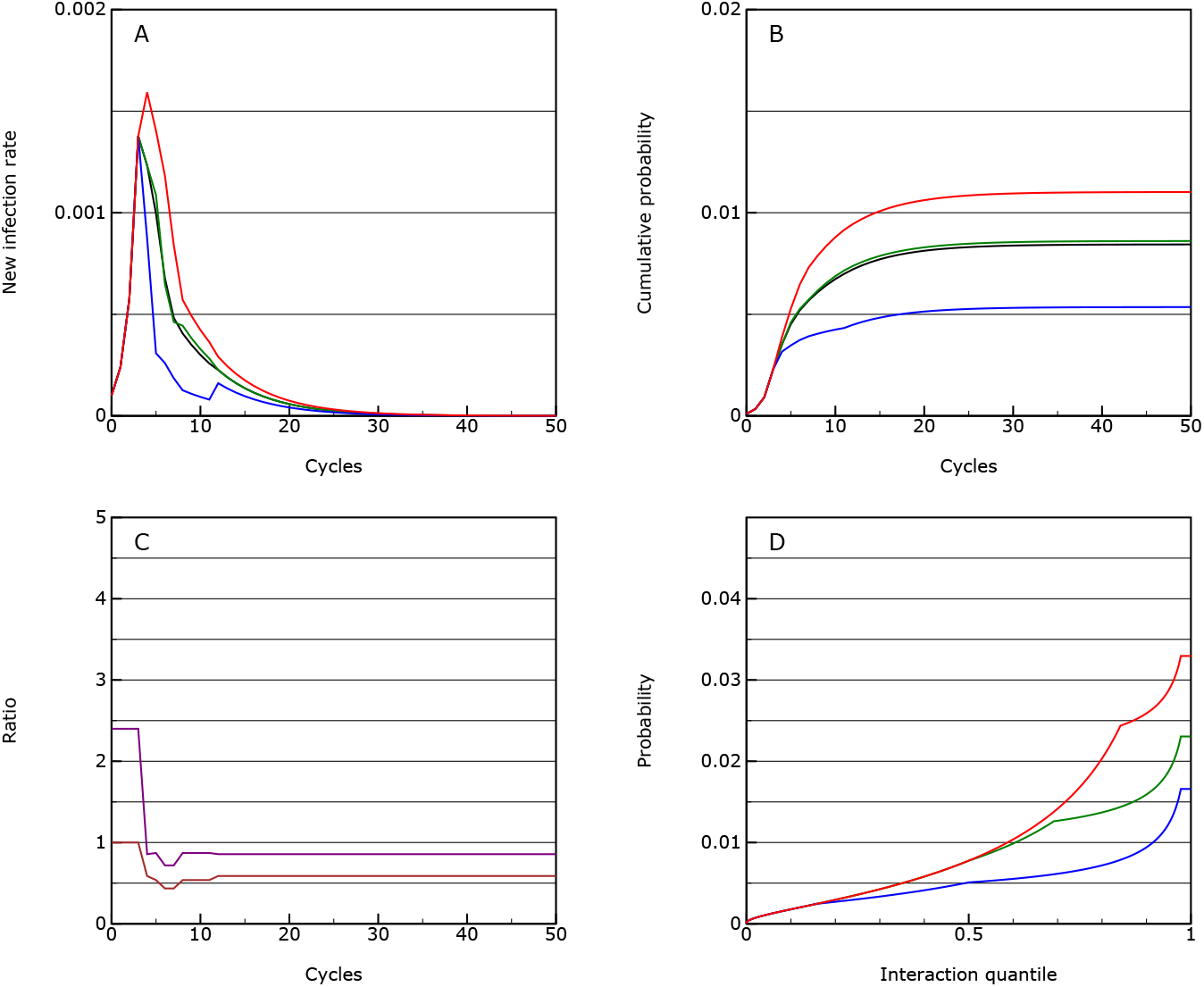
As for Figure 4, but with with measures being imposed continuously until eradication.

The overall activity levels for the 50 cycles are 0.60 of normal levels (or 0.37 if a factor approach is used). However, the advantage here is that after those 50 cycles, potentially all restrictions could be removed.

In this scenario it takes 17 cycles to return to the seed probability. This implies that if you were able to detect an infection rate of 1 in a million within the population and implemented the sanctions immediately, it would take about 17 weeks to eradicate the infection again. This shows that it would certainly be preferable to use good testing and contact tracing methods[5] to prevent reinfection instead.

## 5 Implications of scenario modelling

The scenario tests performed with this model do not throw up any major surprises. The protection scenario is better than the no-intervention approach, but can probably only reduce the mortality rate by a factor of about two. These options would only be justified if the societal costs of other interventions were very high.

Of the other scenarios, the societal cost in terms of lost interaction time all seem to be very similar. For everything but the eradication scenario, it is necessary to have a measured switching on and off of restrictions which keep *R*_eff_ above and below 1. This will be more stable if the changes between the two states is relatively small, both in terms of controlling the epidemic, and in terms of minimising disruption.

The difficulties of control make the eradication scenario particularly attractive. This allows society to adjust to a stable regime over about 50 cycles while the infection rate falls to zero. Once this has been achieved it would then be essential to prevent reinfection through a robust testing and contact tracing mechanism. It is also very likely that such mechanisms could be used at the end of the eradication process to speed up the ending of restrictions.

## 6 Conclusions

For air-borne virus infections, the strong relationship between the probability of picking up an infection and passing it on is critical to evaluation of distancing strategies. In order properly to take account of this it is necessary to consider the very wide range of levels of interaction that individuals have through choices they make, and the jobs that they do.

A very simple model of infection propagation demonstrates that it is much more efficient, when considering interventions to reduce the growth rate of an epidemic to below 1, to target interventions at those activities that result in the very highest interaction levels, and specifically for people who are engaged in those on a regular basis. There is a non-linearity in the system which implies that limiting upper levels of interaction will work more effectively than reducing interactions across the population as a whole. If we can equate lost interaction time with societal cost there is something like a 50% reduction in the cost in taking this approach.

The other consequence of this is that it should be possible to reduce the key *R*0 factor from a value of 2.4, to an an effective value, *R*_eff_, of well below 1, without having a major impact on the majority of the population, if the interventions are properly targeted. Even used on its own, it looks as if sustainable social distancing might be able to eradicate the virus, and this will only be made more practical by other approaches such as testing, quarantining and contact tracing. On the other hand social distancing is not likely to be a good mechanism on its own for preventing re-infection, here testing and contact-tracing[5] would be much more efficient.

Another important implication of this model is that isolation of the majority of the population, while maintaining normal interaction levels for a minority of key workers is likely to result in very high infection rates in that group, and a failure to eradicate the infection within a reasonable time frame. All groups in society must engage in some social distancing or have other protective measures to gain effective control.

The model presented here is a purely theoretical one, based on estimated parameters. It is intended to stimulate further modelling and data collection work which can be used to test the conclusions reached here before they are used to inform policy. However, it is certainly safe to conclude that to reduce *R*_eff_ it is very important to reduce the interaction levels of the most interactive individuals. This is also something seen in simulation and network-based models, but the very simple probability-based model here serves to illustrate why this is case.

## 7 Supplementary information

The model used for the scenario testing is included in the supplementary file Model.html.

## Data Availability

The model is available to be freely distributed.

## References

[1] N. Ferguson, D. Laydon, G. Nedjati Gilani, N. Imai, K. Ainslie, M. Baguelin, S. Bhatia, A. Boonyasiri, Z. Cucunuba Perez, G. Cuomo-Dannenburg, et al., Report 9: Impact of non-pharmaceutical interventions (NPIs) to reduce COVID19 mortality and healthcare demand, Tech. rep., Imperial College London (2020). doi:10.25561/77482. URL http://hdl.handle.net/10044/1/77482

[2] N. M. Ferguson, D. A. T. Cummings, C. Fraser, J. C. Cajka, P. C. Cooley, D. S. Burke, Strategies for mitigating an influenza pandemic, Nature 442 (7101) (2006) 448–452. doi:10.1038/nature04795. URL https://dx.doi.org/10.1038/nature04795

[3] R. Pastor-Satorras, C. Castellano, P. Van Mieghem, A. Vespignani, Epidemic processes in complex networks, Rev. Mod. Phys. 87 (2015) 925–979. doi:10.1103/RevModPhys.87.925. URL https://link.aps.org/doi/10.1103/RevModPhys.87.925

[4] J. Mossong, N. Hens, M. Jit, P. Beutels, K. Auranen, R. Mikolajczyk, M. Massari, S. Salmaso, G. S. Tomba, J. Wallinga, J. Heijne, M. Sadkowska-Todys, M. Rosinska, W. J. Edmunds, Social Contacts and Mixing Patterns Relevant to the Spread of Infectious Diseases, PLoS Medicine 5 (3) (2008) e74. doi:10.1371/journal.pmed.0050074. URL https://dx.doi.org/10.1371/journal.pmed.0050074

[5] L. Ferretti, C. Wymant, M. Kendall, L. Zhao, A. Nurtay, L. Abeler-Dörner, M. Parker, D. Bonsall, C. Fraser, Quantifying SARS-CoV-2 transmission suggests epidemic control with digital contact tracing, Science (2020). doi:10.1126/science.abb6936.

[6] R. Verity, L. C. Okell, I. Dorigatti, P. Winskill, C. Whittaker, N. Imai, G. Cuomo-Dannenburg, H. Thompson, P. Walker, H. Fu, A. Dighe, J. Griffin, A. Cori, M. Baguelin, S. Bhatia, A. Boonyasiri, Z. M. Cucunuba, R. Fitzjohn, K. A. M. Gaythorpe, W. Green, A. Hamlet, W. Hinsley, D. Laydon, G. Nedjati-Gilani, S. Riley, S. van Elsand, E. Volz, H. Wang, Y. Wang, X. Xi, C. Donnelly, A. Ghani, N. Ferguson, Estimates of the severity of COVID-19 disease, medRxiv (2020). doi:10.1101/2020.03.09.20033357.

